# The role of lighting type in dental photography for tooth shade assessment

**DOI:** 10.1101/2025.05.14.25327573

**Authors:** Leszek Szalewski, Dorota Wójcik, Oskar Tokarczuk, Tuana Ozdas, Gabriela Durlej

## Abstract

**Introduction:** Accurate tooth shade assessment is fundamental for achieving optimal aesthetic outcomes in restorative dentistry. This study aims to evaluate the impact of different lighting setups on the reliability and consistency of dental photography for shade selection.

**Materials and Methods:** Three distinct lighting conditions were employed to capture standardized dental photographs: bare twin flash, twin flash with softbox, and twin flash with a polarizing filter. Shade assessments were performed through digital analysis of the photographs and were compared with reference measurements obtained using a spectrophotometer.

**Results:** Preliminary findings indicate that the twin flash with a polarizing filter provides the most accurate tooth color reproduction. However, statistically significant differences were observed when compared to the spectrophotometer measurements. Notably, the closest match to the spectrophotometric values was achieved with light shades (A1, B1) photographed against a black background.

**Conclusions:** The choice of lighting setup significantly affects the accuracy of tooth shade assessment in dental photography. Among the tested configurations, the use of a polarizing filter combined with a black background demonstrates the highest fidelity in color reproduction, particularly for lighter shades.

## Introduction

The color of teeth plays a pivotal role for both patients aspiring to enhance the aesthetic appeal of their smiles and dental professionals striving for precise color matching in restorative procedures [1]. A thorough understanding of color-related phenomena enables clinicians to achieve superior therapeutic outcomes.

Historically, the concept of color in dentistry was viewed as complex, challenging to interpret, evaluate, communicate, and replicate. This perspective shifted in 1904 when Munsell introduced a systematic color notation framework, establishing a three-dimensional color space characterised by lightness (value, the brightness of a color), hue (the perceived color, such as red or green), and chroma (the color’s saturation or purity) [2]. Munsell’s model assigned numerical values to these attributes and used physical reference samples to assess the color of objects [3]. Subsequently, in 1931, the International Commission on Illumination (CIE) established universal standards for color measurement [4]. These standards utilized three coordinates to define colour within a color space, based on systematic consideration of the light source, the observed object, and the observer [5].

The human eye, capable of discerning subtle differences between colors in side-by-side comparisons [6], achieves color perception through three types of cone cells, each tuned to specific wavelength ranges of visible light: short wavelengths (380–500 nm, blue), medium wavelengths (430–600 nm, green), and long wavelengths (530–700 nm, red). The CIE introduced the concept of a “standard observer,” representing an average human’s sensitivity to light across the visible spectrum, effectively linking color perception to specific wavelength responses (XYZ is as an analogue of RGB). The tristimulus values (XYZ) of an object are calculated by multiplying the object’s spectral reflectance (representing reflected light wavelengths) by the spectral distribution of a standard illuminant (derived from the light source’s relative spectral power) and the standard observer’s tristimulus values (XYZ) [4]. This approach has led to the development of instruments such as tristimulus cameras and colorimeters, which utilize transmittance spectra to replicate color coordinates. These devices employ photodetectors to analyze the visible spectrum in digital imaging systems [7].

In 1940, Beckman et al. developed a spectrophotometer capable of analyzing the reflection and transmission properties of objects as a function of wavelength. This advancement provided precise data from the spectral curves, allowing for accurate quantification of CIE color attributes [8]. Over the decades, spectrophotometers have become integral tools for colour evaluation in dentistry, offering high accuracy and reliability [9]. These instruments measure the intensity of light reflected from an object at intervals ranging from 1 to 25 nanometers across the visible spectrum [10].

A spectrophotometer includes a light source, a dispersion mechanism, an optical measuring system, a detector, and a system to convert light data into analyzable signals. In clinical dentistry, spectrophotometric data is processed and interpreted to align with the shade guide standards, facilitating conversion to the equivalent shade tabs [11]. Compared to subjective visual assessments or traditional methods, spectrophotometers have been shown to improve color match accuracy by 33%, achieving objective matches in 93.3% of cases [12].

In contemporary dental practice, digital photography has become an indispensable tool, extensively used for case documentation and communication [13]. Consumer-grade digital and video cameras capture visual data using red, green, and blue light sensors, which are then processed to create a full-color image. Although digital cameras offer a simplified means of electronic shade matching, they still rely on subjective interpretation by the human eye to some extent [14]. High-quality digital images enable precise shade matching by allowing evaluation of multiple areas of a tooth, particularly when paired with appropriate imaging software. This approach is advantageous due to its objectivity, affordability, convenience, ability to evaluate multiple points, streamlined patient record management, and enhanced communication with dental laboratories [13]. To capture optimal images of teeth, smiles, and surrounding soft tissues, facilitating more accurate diagnoses, better color matching, and identification of fine diagnostic details, dental photography often incorporates accessories such as specialized lamps, contrastors, polarizing filters, retractors, and tripods [15]. Furthermore, digital images are easily stored and transmitted electronically, streamlining consultations and communication. Despite these benefits, dental photography does have limitations. The quality of images is highly influenced by the specifications of the camera and lens, while challenges such as color casting must be meticulously managed to ensure precision [13].

### Aim of the study

The main objective of the study was to evaluate the accuracy of reproducing the actual color of the anterior teeth using dental photography, compared to the results obtained with a spectrophotometer. The secondary objective of the study was to evaluate whether there are specific conditions for dental photography that improve the likelihood of achieving clinically acceptable tooth color reproduction, as assessed by the ΔE 2000 parameter, in comparison to the actual color.

## Materials and methods

The study used a Nikon D750 camera equipped with a Nikkor FX 105mm micro 2.8 lens and Godox MF12 K2 twin flash units. All photographs were captured in a darkened room with ambient lighting turned off, employing modelling lamps integrated into flash units for focussing purposes. A constant magnification ratio of 1:1.4 was maintained, corresponding to a distance of 19 cm from the sample. White balance calibration was achieved using a grey card. Exposure settings remained consistent across all images: aperture f/25, ISO 100, and shutter speed of 1/125 s. The flash power output was adjusted according to the light modifiers used: 1/8 power for flashes without modifiers, 1/4 power with FixLite softboxes, and full power when polarizing filters (Intra.Photos) were applied. Half of the photos were taken with a tripod, and the other half were taken handheld by the operator. Color measurements of the captured images were performed in the CIELab color space using ColorChecker software on a MacBook Pro.

A standard VITA shade guide containing 10 shades (A1, A2, A3, A4, B1, B2, C3, C4, D3 and D4) was used in the study. The selected shades represent the full spectrum of human tooth colouration, ranging from very light (B1) to very dark (C4, D4).

Reference measurements of the standard samples were conducted using a VITA MASTER spectrophotometer in the CIELab color space. The values obtained in this system were subsequently converted to their corresponding dental shades (A1, A2, A3, etc.). Reference values in the CIELab system were established as follows: L = 81.0; a = 2.3; b = 20.0.

Subsequently, dental photographs of each tooth shade were taken under various combinations of lighting conditions, backgrounds, and photographic techniques, resulting in a total of 1,200 measurements. The lighting variable was assessed under three conditions: twin flash units without light modifiers (TF group), twin flash units with FixLite softboxes (SB group), and twin flash units with polarizing filters (FP group). Background variations included two types: black and white, while the exposure variable encompassed two photography techniques: tripod-mounted and handheld shots. Each shade from the VITA shade guide was photographed 10 times under each combination of lighting, background, and exposure method, yielding 120 measurements for each shade. Color difference assessments (ΔE) between reference measurements and the values obtained from dental photographs were conducted based on formulas (1) and (2), reflecting older and more contemporary standards [16].

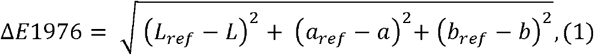

where:

- (*L*_*ref*_ − *L*) represents the difference in lightness between e reference color and the measured value, Δ*L*′
- (*a*_*ref*_ − *a*) denotes the difference on the red-green axis,
- (*b*_*ref*_ − *b*) denotes the difference on the yellow-blue axis.

Formula 2 represents a more recent standard that incorporates additional parameters that affect the perception of color differences by the human eye, such as scaling functions and rotation coefficients. Due to its complexity, this formula requires the use of computer algorithms for calculations:

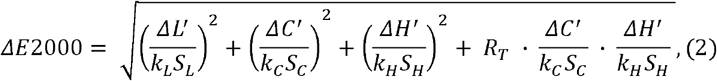

where:

- Δ*L*′-lightness difference between the reference color and the measured color
- Δ*C*′-chroma difference between the reference color and the measured color,
- Δ*H*′-hue difference between the reference color and the measured color,
- *k*_*L*_, *k*_*C*_, *k*_*H*_ – rotation coefficient.
- *S*_*L*_, *S*_*C*_, *S*_*H*_ – scaling coefficient,
- *R*_*T*_ – scaling functions for lightness, chroma and hue,

The ΔE results were interpreted based on the scale presented in Table 1.

**Table 1.** Interpretation of ΔE results [16].

| $\Delta E$ 1976/2000 | Result |
| --- | --- |
| 0-2 | Difference perceptible only to an experienced observer |
| 2-5 | Difference noticeable to an inexperienced observer or a clear color difference |
| >5 | Noticeable color difference, classified as two distinct colors. |

### Statistical analysis

The significance level of the statistical tests was set at α = 0.05.

The distribution of numerical variables was characterized using descriptive statistics. For this purpose, measures of central tendency, represented by the mean, and measures of dispersion, represented by the standard deviation (SD), were employed. Categorical variables were described using frequency counts.

### Bland Altman analysis

To evaluate the agreement between the two methods of measuring color differences (ΔE 2000 and ΔE 1976), the Bland-Altman method was applied [17].

### Mean delta L, delta C, delta H

In this study, Formula 2 was applied to evaluate color differences, incorporating additional standardization of the results for assessing color discrepancies. Perceptual thresholds for the Formula 2 parameters were adopted according to Ghinea et al. [18], defined as the “threshold 50/50,” representing the boundary values at which 50% of observers perceive a difference between two colors. The threshold values were established as follows: L′ < |2.44| for lightness, ΔC′ < |3.15| for chroma, and ΔH′ < |3.24| for hue.

Values below these thresholds were considered clinically acceptable. A desired clinical outcome was defined as achieving a total color difference of ΔE 2000 < 2, indicating a difference that could only be perceived by an experienced observer.

### Firth Logistic regression

To estimate the magnitude and direction of the effects of individual parameters involved in dental photography on achieving the desired clinical outcome (ΔE 2000 < 2), the Firth logistic regression model [19] was applied. We selected Firth’s logistic regression model due to the low frequency of the target outcome, which occurred in only 17 out of 1,200 observations, representing a prevalence of approximately 1.4%. The full model specification is provided in the Supplement.

Model fit was assessed using the likelihood ratio test (LRT) and the Wald z-test, while the explained variance was determined by McFadden’s R^2^ coefficient. Effect sizes were expressed as odds ratios (OR) along with 95% confidence intervals, which were estimated using the profile likelihood method. The 95% CI and p-values were evaluated using the asymptotic approximation of the Wald z-test statistics.

### Statistical environment

Analyses were conducted using the R Statistical language (version 4.4.1; R Core Team, 2024) on Windows 11 pro 64 bit, using the packages: *ggstatsplot* (version 0.9.3)[20], *ggplot2* (version 3.4.0) [21], *readxl* (version 1.3.1) [22] and *dplyr* (version 1.1.2) [23], *effectsize* (version 0.8.3) [24] and *psych* (version 2.1.6) [25], *sjPlot* [26], *logistf* [27], *viridis* [28], *ggrepel* [29], *colorspace* [30].

## Results

Descriptive Statistics

Reference Values

The reference values were obtained as the average of four measurements taken with a spectrophotometer for each shade, and their summary is presented in Table 2.

**Table 2.** Reference values for L, a, b obtained from spectrophotometric measurements of VITA shade guide samples.

| Shade | Reference L | Reference a | Reference b |
| --- | --- | --- | --- |
| A1 | 81,5 | 2,3 | 19,7 |
| A2 | 76,2 | 0,1 | 23,6 |
| A3 | 75,8 | 0,2 | 26,7 |
| A4 | 67,0 | 4,9 | 31,8 |
| B1 | 84,6 | -2,8 | 20,2 |
| B2 | 80,0 | -1,8 | 24,8 |
| C3 | 72,8 | 0,4 | 28,1 |
| C4 | 64,3 | 4,2 | 30,7 |
| D3 | 70,7 | 2,5 | 25,5 |
| D4 | 72,7 | -2,3 | 27,8 |

### Mean and Standard Deviation of Delta E 2000

The lowest Delta E 1976 values (0.62) were observed for the A1 shade photographed with a polarizing filter on a tripod against a black background. All clinically significant results (ΔE 2000 < 2) were achieved with a polarizing filter on a tripod against a black background (0.49 - 1.17) and handheld (1.67 - 1.84).

For the ΔE 2000 results, the lowest values were obtained for the A1 shade photographed on a black background with a polarizing filter on a tripod (0.42 - 0.91). Clinically significant results were obtained for the A1 shade with a polarizing filter on a black background (1.73 - 1.87). The lowest mean value of ΔE 2000 was observed for the A1 shade and photographs taken on a black background with a polarizing filter on a tripod (0.55) (Table 3).

**Table 3.**
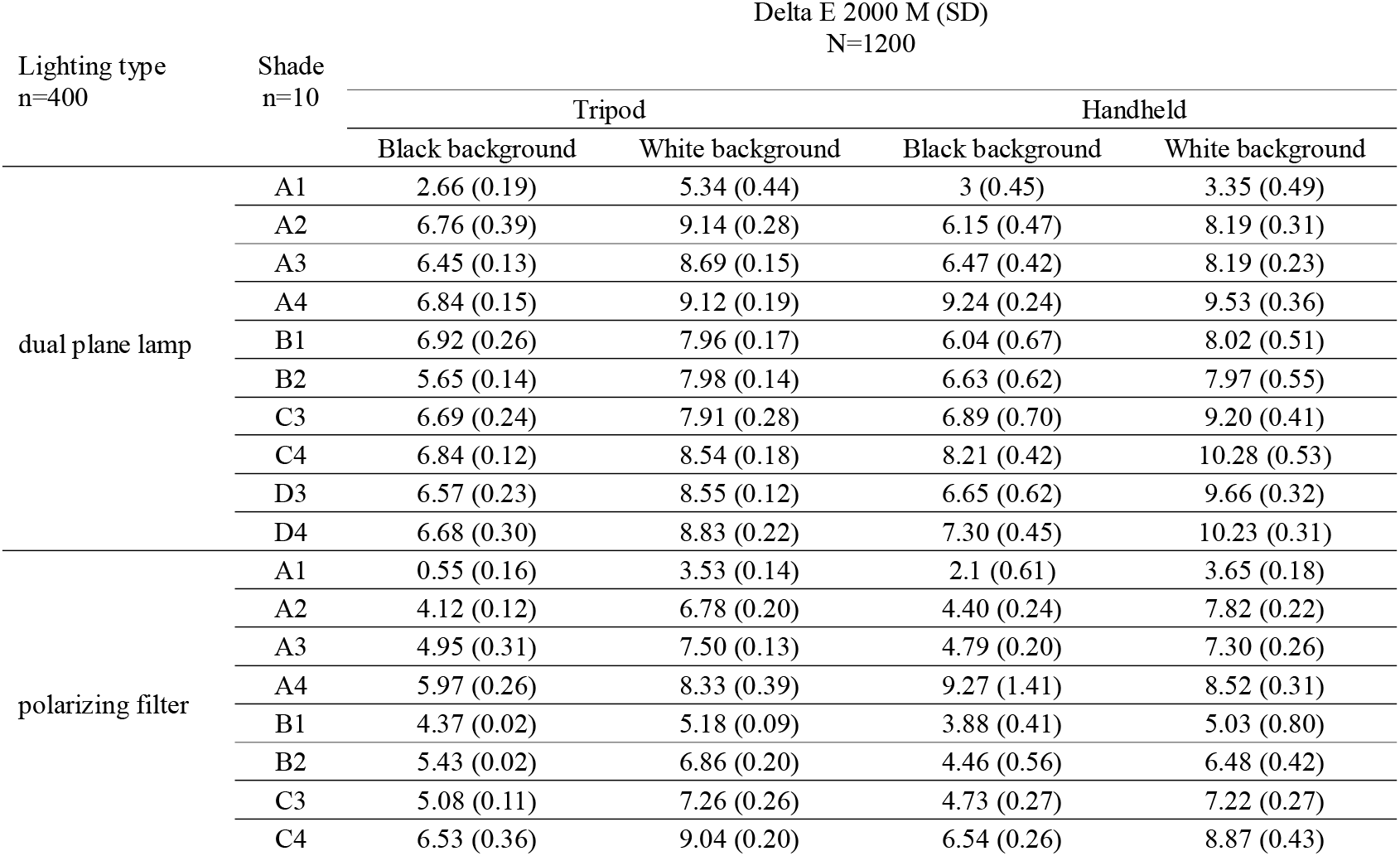
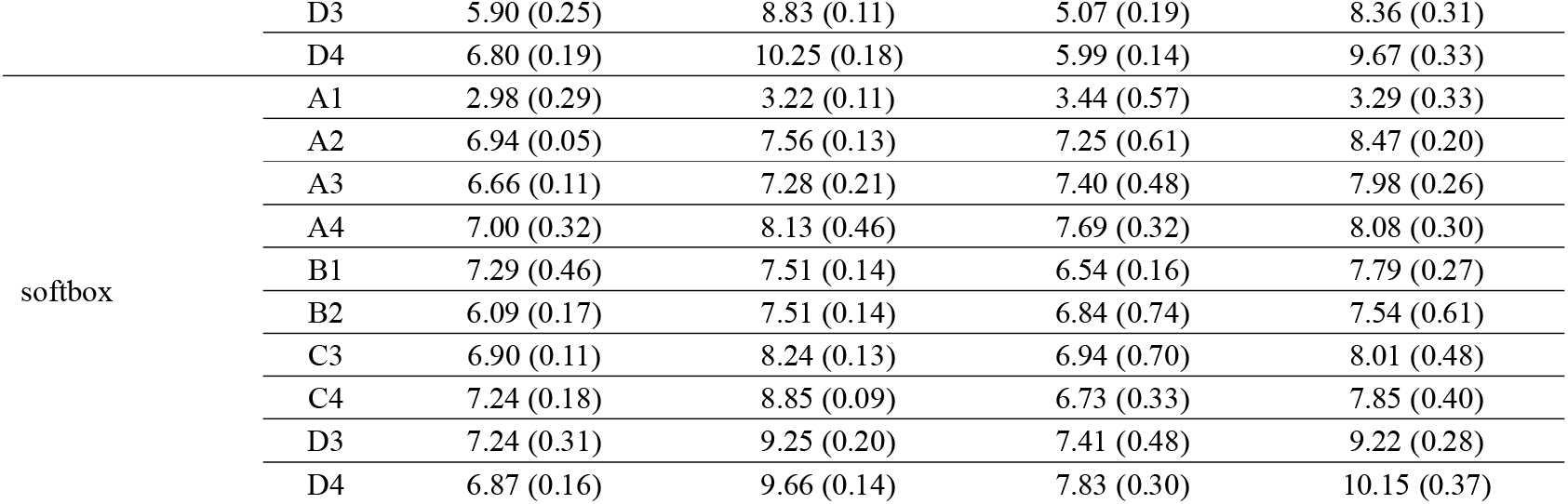
Mean and SD for ΔE 2000 categorized by photographic method, background color, and type of flash.

### Mean ΔL, ΔC, and ΔH

The aim of the analysis was to assess the parameters of lightness (|ΔL′| < 2.44), chroma (|ΔC′| < 3.15), and hue (|ΔH′| < 3.24). Clinically significant cut-off points were based on the results obtained by Ghinea (threshold 50/50), which define the boundary for the acceptability of color differences.

Lightness Difference (ΔL′): Positive values indicate a lighter sample compared to the reference, while negative values indicate a darker sample.

Chroma Difference (ΔC′): Positive values signify a higher colour saturation of the sample relative to the reference, while negative values indicate lower saturation.

Hue Difference (ΔH′): Positive values indicate a hue shift in the clockwise direction in the ab plane of the CIELAB color space, while negative values indicate a shift in the counterclockwise direction.

### Delta L

The analysis revealed that only a subset of the photographic parameter combinations met the clinical criterion for acceptability regarding lightness (|ΔL′| < 2.44). It was observed that better lightness reproduction was achieved in photos taken against a black background compared to a white background, both when using a tripod and handheld (detailed results are provided in Table 3 in the supplement). The highest values of |ΔL′|, exceeding the threshold of 2.44, were observed for photos taken against a white background using a tripod with a polarizing filter (A4, C3, D3, D4 colors) and a softbox (D3, D4 colors). Among the tested colors, only A4 was lighter than the reference, while the others exhibited a tendency to darken. The largest deviations in ΔL′ towards darkening were observed on a white background for dark colors with dual flash (A4, C3, C4, D3, D4) and polarizing filter (C4, D3, D4).

### Delta C

Clinically acceptable differences in color saturation were more frequently observed in photographs taken against a black background, regardless of whether a tripod was used or the camera was handheld. The worst color saturation reproduction occurred for the B1 color photographed with dual-plane lighting on a white background with a tripod and the D4 color photographed using a polarizing filter on a white background handheld.

### Delta H

The analysis of hue reproduction indicated that ΔH′ values fell below the clinical acceptability threshold (|ΔH′| < 3.24) only for the A1 shade, regardless of the combination of lighting, background, and photographic method. The largest ΔH′ deviations were observed for handheld photos on a white background with dual flash (A2, A3, B2, C3-C4, D3-D4 colors) and softbox (A2-D4 colors), as well as on a black background with handheld softbox (A4, C4, D4 colors). For tripod-based photography, significant ΔH′ deviations occurred on a white background with dual flash (A2, A3, B2-D4 colors) and softbox (A2-A3, C3, C4, D4 colors), and on a black background with softbox (A2-A3, C3-D4 colors). All observed ΔH′ values were positive, indicating a hue shift in the clockwise direction on the color wheel relative to the reference color.

The results suggest that clinically acceptable colour reproduction in terms of lightness, saturation, and hue is achievable, particularly for the A1 color when using a polarizing filter, regardless of the background color or camera stabilization method employed.

The Bland Altman analysis results in Figure 1 revealed a systematic bias, indicating that ΔE1976 consistently produces higher values than ΔE2000. Bias 1.73, UL – 0,28 and LL -3,74. (Table 4). Most points fall within the agreement range. For high ΔE 2000 values, starting from approximately 9, the differences between the methods exceed the lower limit of agreement, indicating larger discrepancies for the shades C4, D3, and D4. Both methods, except for the colours C4, D3, and D4, can be used interchangeably.

**Table 4.**
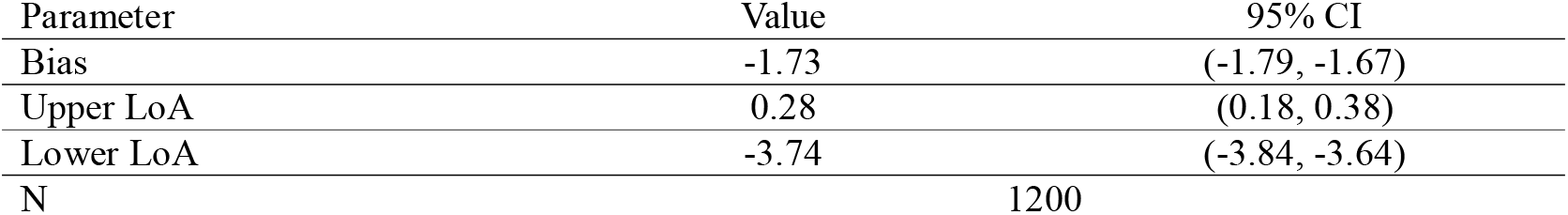
Summary of Bland-Altman analysis for ΔE1976 vs ΔE2000 comparison Parameter Value 95% CI.

| Parameter | Value | 95% CI |
| --- | --- | --- |
| Bias | -1.73 | (-1.79, -1.67) |
| Upper LoA | 0.28 | (0.18, 0.38) |
| Lower LoA | -3.74 | (-3.84, -3.64) |
| N | 1200 |  |

**Figure 1.**
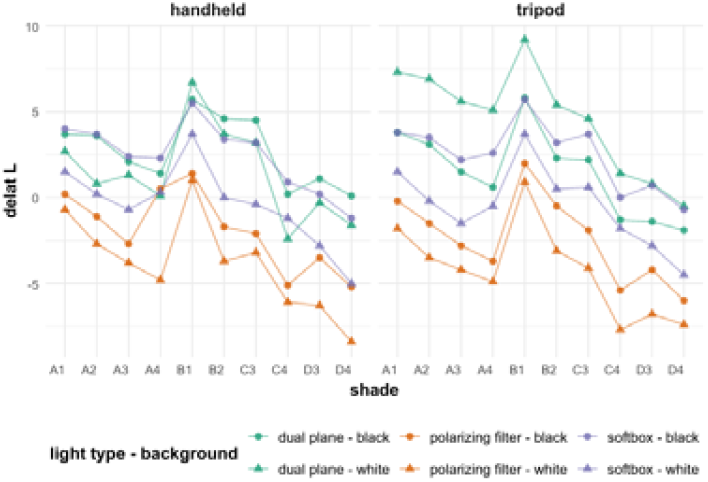
Average ΔL′, values under different photographic conditions for analyzed shades.

**Figure 2.**
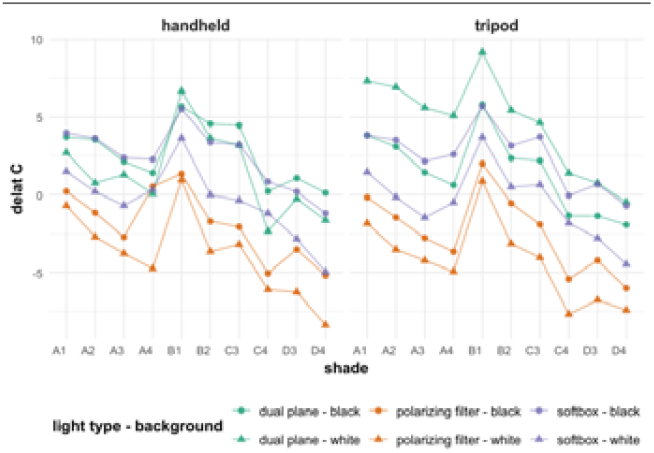
Average ΔC′ values under different photographic conditions for analyzed shades.

**Figure 3.**
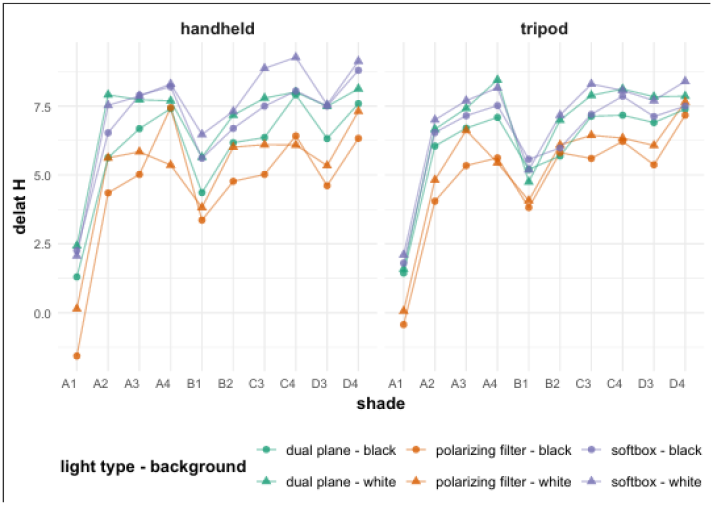
Average ΔH′ values under different photographic conditions for analyzed shades.

**Figure 4.**
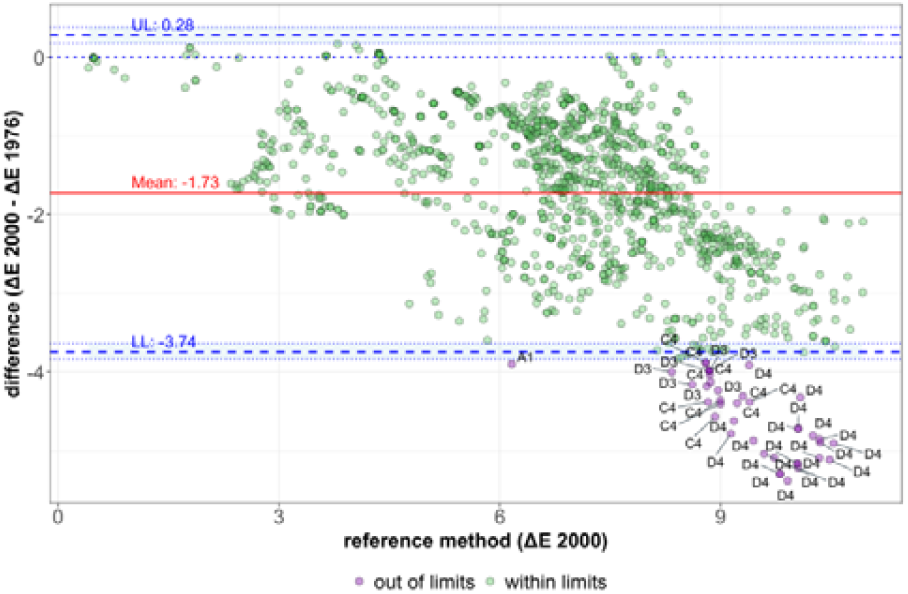
Bland-Altman Plot: agreement between ΔE 2000 and ΔE 1976.

**Figure 5.**
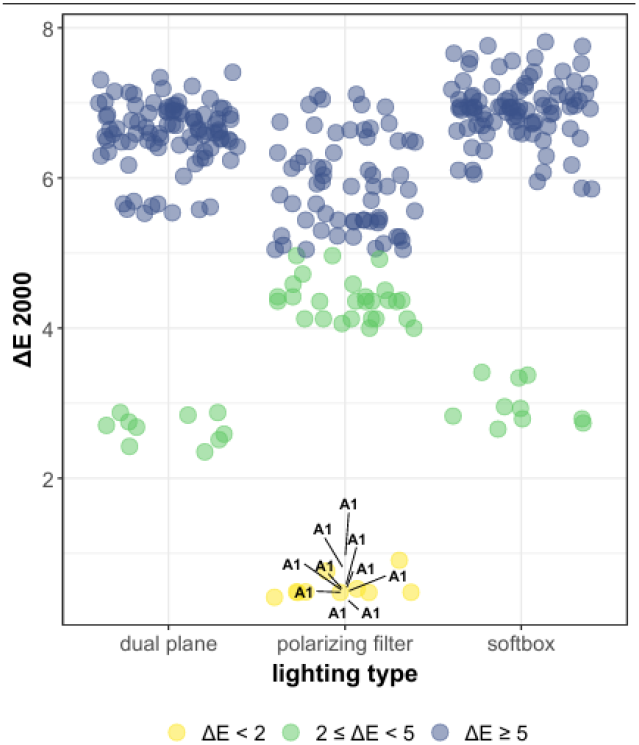
ΔE 2000 values by photographic conditions on black backdrop - tripod mounted camera.

**Figure 6.**
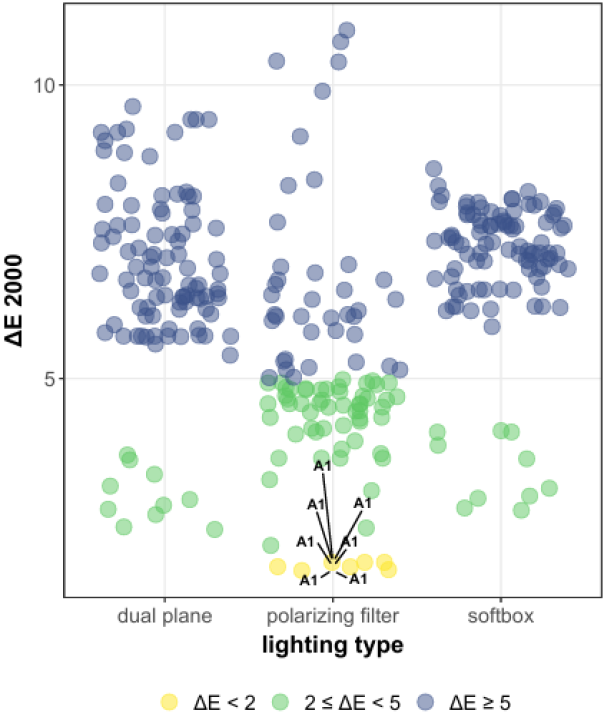
ΔE 2000 values by photographic conditions on black backdrop - handheld camera.

**Figure 7.**
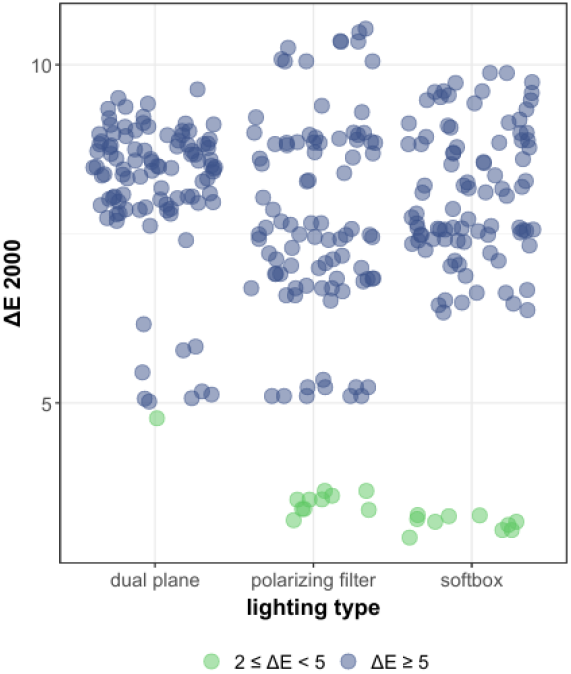
ΔE 2000 values by shade and photographic conditions on white backdrop - tripod mounted camera.

**Figure 8.**
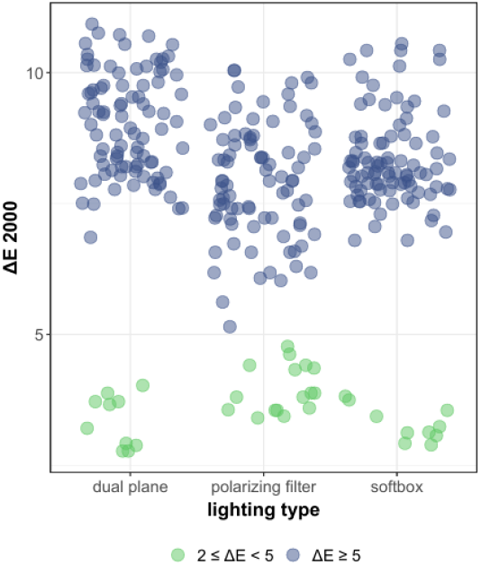
ΔE 2000 values by shade and photographic conditions on white backdrop - handheld camera.

### Analysis of ΔE 2000 values by shade and type of photograph

The analysis of the ΔE 2000 results, categorized by background type and photographic conditions, reveals noticeable differences in the distribution of values. Clinically acceptable color differences (ΔE 2000 < 2), distinguishable only by a trained observer, were observed exclusively on a black background, both when using a tripod and when the camera was handheld, with the application of a polarizing filter. The differences in the obtained results may be attributed to variations in the angle of the photographs, depending on how the operator holds the camera. Slightly greater differences were observed in images taken handheld, compared to those taken with a tripod, due to varying angles of camera positioning. For the black background, the desired effect was achieved. However, for a white background, no target outcome was achieved for any combination of lighting and photographic method, indicating that a white background may significantly reduce the chances of obtaining the desired color reproduction in photographs. The target effect (ΔE 2000 < 2) was achieved in 17 measurements (1.42%). Clearly distinguishable color differences (ΔE 2000 ≥ 5) were observed in 1005 cases (83.75%). The remaining measurements (178, 14.83%) fell within the range of 2 ≤ ΔE 2000 < 5.

ΔE 2000 values ≥ 5 were significantly more frequent on a white background, especially when using a softbox or dual-plane lighting, suggesting greater challenges in achieving accurate color reproduction under these conditions.

For the analysis, a binary variable ΔE binary was created, where the desired outcome (favorable outcome) was defined as ΔE < 2. This coding is based on the ΔE classification, according to which the values below 2 indicate that the color difference between the spectrophotometric measurement and the photograph is imperceptible to the average observer:

Code 1: ΔE 2000 < 2 (color difference perceptible only to a trained observer)

Code 0: ΔE 2000 ≥ 2

### Model

The reference category for the model is dual plane as the light type, handheld as the photo type, and white as the backdrop. The odds ratio (OR = 0.0021, 95% CI 0.0001–0.033, p < 0.001) indicates that, under these conditions, the probability of achieving an acceptable color difference is very low. According to the results in Table 5, the use of a polarizing filter increases the likelihood of obtaining an acceptable color difference by more than 38 times (OR = 38.15, 95% CI [2.03, 488.22], p < 0.001). A black background also increases these odds by 38 times (OR = 0.03, 95% CI [2.54 – 572.97], p < 0.001). The impact of tripod stabilization, although suggesting a positive trend (OR = 1.45, 95% CI [0.57, 3.70]), did not reach statistical significance (p = 0.460). The use of a soft box did not show a significant impact on achieving an acceptable color difference compared to the dual flash lamp (OR = 1.00, 95% CI [0.02, 45.71], p = 1.000). A McFadden’s R^2^ of 0.315 indicated that the model exhibited a satisfactory fit to the data. Both the polarizing filter and the black background contributed equally to achieving clinically acceptable ΔE values, essential for accurate color reproduction in dental photography. The magnitude of the effects in both cases is large, but they should be interpreted cautiously due to the wide confidence intervals. The results underscore the importance of optimal lighting and background selection for precise photographic documentation in dentistry.

**Table 5.** Logistic regression results using Firth’s method.

| Predictors | OR | 95% CI | *p* |
| --- | --- | --- | --- |
| (Intercept) | $1.0 \cdot 10^{-4}$ | $1.00 \cdot 10^{-6} - 2.50 \cdot 10^{-3}$ | **< 0.001** |
| light type [polarising filter] | 38.15 | 2.45 – 593.66 | < **0.001** |
| light type [softbox] | 1.00 | 0.02 – 45.71 | 1.000 |
| photo type [tripod] | 1.45 | 0.57 – 3.70 | 0.460 |
| backdrop [black] | 38.13 | 2.54 – 572.97 | < **0.001** |
| Observations | 1200 |  |  |
| LRT | $\chi^2 (4) = 53.55, p < 0.001$ | | |
| Wald test | $\chi^2 (4) = 144.78, p < 0.001$ | | |
| R<sup>2</sup> McFadden: | 0.315 |  |  |

The model explains approximately 31.5% of the variability in the analyzed data, suggesting that there are many other factors that may affect the variability of the results, including those that cannot be controlled. Future studies should consider additional factors that could impact measurement quality. A critical element is the angle of photographing the object, which is directly related to the distance from the photographed object. Equally important could be the angle of light incidence or the intensity of lighting used during the photography. The results obtained support the main hypothesis.

## Discussion

The results obtained indicate that the colors achieved through color measurement according to the CIELAB scale differ from those obtained using spectrophotometric measurements, regardless of the lighting conditions or the background used. Analysis of the individual color components, lightness, hue, and chroma—revealed that the best color reproduction was achieved for the lightness component.

Considering the complexity of the colour of human tooth color, the study utilized only a small fraction of the possible color combinations. Hein et al. conducted a color analysis of 8,153 natural teeth and identified as many as 1,173 unique tooth shades [31]. Thus, relying solely on traditional dental color systems, such as the Vita Master scale with its 26 shades, may be insufficient, and alternative methods should be explored. One potential solution is the use of dental photography with polarising filters, a concept introduced by Hein et al. [32]. However, studies have shown that the application of polarizing filters and a black background did not allow for perfect color reproduction in photographs when compared to spectrophotometer measurements. Mahn et al. obtained similar findings in their research comparing color matching using a spectrophotometer and photography with polarizing filters [33]. Consistent with our findings, the most accurate results were obtained for the L (lightness) component. Nevertheless, compared to visual color assessment with a shade guide, significantly more accurate results were achieved with photographic methods. Consequently, the authors suggest that photography could be a superior alternative to visual evaluation. Additionally, Sirintawat et al. demonstrated that photography, regardless of the camera type - DSLR or smartphone, and irrespective of the use of polarizing filters, did not replicate colors as accurately as a spectrophotometer [38]. Delta E values ranged from 6 for intraoral scanners to 20 for photographs taken with a polarizing filter. However, these findings were limited by the absence of information on whether a gray card was used for post-production. Without a grey card, photography with a polarizing filter is unlikely to be effective. These results are in line with a systematic review from 2012 [34], which analyzed 26 studies and concluded that the most repeatable results are obtained with a spectrophotometer, followed by photography with polarizing filters, and finally, visual assessment.

Visual color evaluation proved to be the most subjective method, potentially explaining its lower accuracy. Similar conclusions were drawn in a more recent systematic review from 2023 [36], which analyzed 85 studies and reaffirmed that the spectrophotometer remains the gold standard. In some studies, dental photography achieved a Delta E below the acceptable threshold (<2) when compared to spectrophotometer results; however, in most studies, the differences exceeded this threshold. The results obtained by visual assessment showed the greatest discrepancies compared to the spectrophotometer measurements. Patamar et al. reported findings similar to ours, demonstrating greater accuracy for lighter shades (A1, B1) compared to darker ones (A3, B3) [37].

Regarding color matching using photography, there is a wide range of available equipment, including DSLRs, mirrorless cameras, as well as full-frame and crop-sensor cameras. Jasim et al. compared color reproduction using the same polarizing filters with both full-frame and crop-sensor cameras [39]. Their findings clearly suggest that full-frame cameras reproduce colors more accurately. In the case of full-frame cameras, the results remained within the acceptable threshold (Delta E <2), while for crop-sensor cameras, the Delta E value was 3.5, exceeding the threshold. In contrast, Farah et al. demonstrated that the type of lighting (LED, fluorescent, or natural light) in the environment where color matching is performed does not significantly affect the results. The findings by Akl et al. challenge the results obtained with dental spectrophotometers when compared with technical spectrophotometers [35]. The differences were statistically significant, highlighting the need to search for alternative methods that could replace the current ones, which present several limitations.

Both factors, the use of a polarizing filter as a light modifier and the application of a black background, play significant and equivalent roles in achieving the target Delta E difference, which allows for accurate color reproduction in dental photography. The effect sizes in both cases are substantial, although they should be interpreted with caution due to wide confidence intervals. These findings underscore the importance of optimizing lighting and background conditions for precise photographic documentation in dentistry. Further research is necessary to identify optimal conditions and accessories for dental photography that may eventually enable its use for ideal tooth color matching.

## Conclusion

Based on the obtained results, it can be concluded that accurate color reproduction of teeth in photographs remains unachievable regardless of the applied lighting conditions or background color. However, the use of polarizing filters combined with a black background enhances color fidelity, particularly in the case of lighter shades.

## Data Availability

All data produced in the present study are available upon reasonable request to the authors

## References

1. Tabatabaian F, Beyabanaki E, Alirezaei P, Epakchi S. Visual and digital tooth shade selection methods, related effective factors and conditions, and their accuracy and precision: A literature review. J Esthet Restor Dent. 2021 Dec;33(8):1084–1104. doi: 10.1111/jerd.12816. Epub 2021 Sep 9. PMID: 34498789.

2. Nickerson D. The Munsell color system. Illum Eng. 1946 Jul;41(7):549–60. PMID: 20285754.

3. Billmeyer FW Jr, Webber AC. Three-dimensional color models constructed on the CIE and Munsell systems. J Opt Soc Am. 1953;43: 69–70.

4. Smith T, Guild J. The C.I.E. colorimetric standards and their use. Trans Opt Soc. 1931;33:73–134.

5. Wyszecki G, Sanders CL. Correlate for lightness in terms of CIEtristimulus values. Ii J Opt Soc Am. 1957;47:840–842.

6. Igiel C, Lehmann KM, Ghinea R, Weyhrauch M, Hangx Y, Scheller H, Paravina RD. Reliability of visual and instrumental color matching. J Esthet Restor Dent. 2017 Sep;29(5):303–308. doi: 10.1111/jerd.12321. Epub 2017 Jul 25. PMID: 28742283.

7. Valberg A. A visual tristimulus projection colorimeter. Appl Optics. 1971;10:8–13.

8. Beckman AO, Gallaway WS, Kaye W, et al. History of spectrophotometry at Beckman instruments Inc. Anal Chem. 1977;49:280A300A.

9. Paul SJ, Peter A, Rodoni L, Pietrobon N. Conventional visual vs spectrophotometric shade taking for porcelain-fused-to-metal crowns: a clinical comparison. Int J Periodontics Restorative Dent. 2004 Jun;24(3):222–31. PMID: 15227770.

10. Khurana R, Tredwin CJ, Weisbloom M, Moles DR. A clinical evaluation of the individual repeatability of three commercially available colour measuring devices. Br Dent J. 2007 Dec 22;203(12):675–80. doi: 10.1038/bdj.2007.1108. PMID: 18084212.

11. Lagouvardos PE, Fougia AG, Diamantopoulou SA, Polyzois GL. Repeatability and interdevice reliability of two portable color selection devices in matching and measuring tooth color. J Prosthet Dent. 2009 Jan;101(1):40–5. doi: 10.1016/S0022-3913(08)60289-9. PMID: 19105990.

12. Paul S, Peter A, Pietrobon N, Hämmerle CH. Visual and spectrophotometric shade analysis of human teeth. J Dent Res. 2002 Aug;81(8):578–82. doi: 10.1177/154405910208100815. PMID: 12147751.

13. Chu SJ, Trushkowsky RD, Paravina RD: Dental color matching instruments and systems. Review of clinical and research aspects. J Dent. 2010, 38 Suppl 2:e2–16. 10.1016/j.jdent.2010.07.001

14. Blaes J. Today’s technology improves the shade-matching problems of yesterday. J Indiana Dent Assoc. 2002-2003 Winter; 81(4):17–9. PMID: 12593181.

15. Singh A, Prasad AB, Raisingani D, Srivastava H, Moryani V. Capturing the art and science of dentistry in a lens: Digital dental photography. J Conserv Dent Endod. 2024 May;27(5):449–457.

16. Sharma, G., Wu, W. and Dalal, E.N. (2005), The CIEDE2000 color-difference formula: Implementation notes, supplementary test data, and mathematical observations. Color Res. Appl., 30: 21–30. 10.1002/col.20070

17. Bland JM, Altman DG. Measuring agreement in method comparison studies. Stat Methods Med Res. 1999;8(2):135–160.

18. Ghinea R, Pérez MM, Herrera LJ, Rivas MJ, Yebra A, Paravina RD. Color difference thresholds in dental ceramics. J Dent. 2010;38 Suppl 2:e57–64. doi: 10.1016/j.jdent.2010.07.008. Epub 2010 Jul 27. PMID: 20670670.

19. Kosmidis, I., & Firth, D. (2021). Jeffreys-prior penalty, finiteness and shrinkage in binomial-response generalized linear models. Biometrika, 108(1), 71–82.

20. Patil, I., (2021). Visualizations with statistical details: The ‘ggstatsplot’ approach. Journal of Open Source Software, 6(61), 3167, 10.21105/joss.03167

21. Wickham H. ggplot2: Elegant Graphics for Data Analysis. 2nd ed. Cham: Springer; 2016.

22. Wickham, H., & Bryan, J. (2019). readxl: Read Excel files [Computer software]. Retrieved from https://cran.r-project.org/web/packages/readxl/index.html

23. Wickham H, François R, Henry L, Müller K, Vaughan D (2023). dplyr: A Grammar of Data Manipulation. R package version 1.1.4, https://github.com/tidyverse/dplyr,

24. Ben-Shachar et al., (2020). effectsize: Estimation of Effect Size Indices and Standardized Parameters. Journal of Open Source Software, 5(56), 2815, 10.21105/joss.02815

25. Revelle, W. (2021a). psychTools: Tools to Accompany the Psych Package for Psychological Research. Northwestern University, Evanston, https://CRAN.r-project.org/package=psychTools. R package version 2.1.6

26. Lüdecke D, Lüdecke MD. Package sjPlot. The Comprehensive R Archive Network. 2015. [01-07-2024]. https://cran.r-project.org/web/packages/sjPlot/index.html

27. Heinze G, Ploner M, Dunkler D, Southworth H. logistf: Firth’s Bias-reduced Logistic Regression. R package, Version 1.23. https://CRAN.R-project.org/package=logistf

28. Garnier, S., Ross, N., Rudis, R., Camargo, P. A., Sciaini, M., & Scherer, C. (2021). viridis-Colorblind-friendly color maps for R. R package version 0.6, 2.

29. Slowikowski K, Schep A, Hughes S, Lukauskas S, Irisson JO, Kamvar ZN, Ryan T, Christophe D, Hiroaki Y, Gramme P. “Package Ggrepel: automatically position non-overlapping text labels with ‘Ggplot2. “. 0.9.4Ggrepel. 2018 https://rdrr.io/cran/ggrepel/

30. Zeileis, A., Fisher, J. C., Hornik, K., Ihaka, R., McWhite, C. D., Murrell, P., … Wilke, C. O. (2020). colorspace: A Toolbox for Manipulating and Assessing Colors and Palettes. Journal of Statistical Software, 96(1), 1–49. 10.18637/jss.v096.i01

31. Hein S, Morovič J, Morovič P, Saleh O, Lüchtenborg J, Westland S. How many tooth colors are there? Dent Mater. 2025 Jan;41(1):51–57. doi: 10.1016/j.dental.2024.10.016. Epub 2024 Oct 29. PMID: 39472195.

32. Hein S, Modrić D, Westland S, Tomeček M. Objective shade matching, communication, and reproduction by combining dental photography and numeric shade quantification. J Esthet Restor Dent. 2021 Jan;33(1):107–117. doi: 10.1111/jerd.12641. Epub 2020 Aug 24. PMID: 32840048.

33. Mahn E, Tortora SC, Olate B, Cacciuttolo F, Kernitsky J, Jorquera G. Comparison of visual analog shade matching, a digital visual method with a cross-polarized light filter, and a spectrophotometer for dental color matching. J Prosthet Dent. 2021 Mar;125(3):511–516. doi: 10.1016/j.prosdent.2020.02.002. Epub 2020 Mar 17. PMID: 32197819.

34. Chen H, Huang J, Dong X, Qian J, He J, Qu X, Lu E. A systematic review of visual and instrumental measurements for tooth shade matching. Quintessence Int. 2012 Sep;43(8):649–59. PMID: 23034418.

35. Akl MA, Sim CPC, Nunn ME, Zeng LL, Hamza TA, Wee AG. Validation of two clinical color measuring instruments for use in dental research. J Dent. 2022 Oct;125:104223. doi: 10.1016/j.jdent.2022.104223. Epub 2022 Jul 15. PMID: 35839964.

36. Rashid F, Farook TH, Dudley J. Digital Shade Matching in Dentistry: A Systematic Review. Dent J (Basel). 2023 Oct 27;11(11):250. doi: 10.3390/dj11110250. PMID: 37999014; PMCID: PMC10670912.

37. Patankar, A.H.; Miyajiwala, J.S.; Kheur, M.G.; Lakha, T. Comparison of photographic and conventional methods for tooth shade selection: A clinical evaluation. J. Indian Prosthodont. Soc. 2017, 17, 273–281

38. Sirintawat N, Leelaratrungruang T, Poovarodom P, Kiattavorncharoen S, Amornsettachai P. The Accuracy and Reliability of Tooth Shade Selection Using Different Instrumental Techniques: An In Vitro Study. Sensors (Basel). 2021 Nov 11;21(22):7490. doi: 10.3390/s21227490. PMID: 34833565; PMCID: PMC8620419.

39. Jasim A, Al-Qazzaz S, Radhi N. Assessment of full frame and crop frame DSLR cameras for dental shade selection: in vitro comparative study. Journal of Stomatology. 2024;77(4):278–284. doi:10.5114/jos.2024.145793.

40. Farah RI, Almershed AS, Albahli BF, Al-Haj Ali SN. Effect of ambient lighting conditions on tooth color quantification in cross-polarized dental photography: A clinical study. J Prosthet Dent. 2022 Oct;128(4):776–783. doi: 10.1016/j.prosdent.2021.01.015. Epub 2021 Feb 25. PMID: 33640092.

